# Multimorbidity and its associated factors in Indonesia through universal health coverage scheme: A cross-sectional study based on national claims data

**DOI:** 10.1101/2022.09.19.22280100

**Authors:** Noor Afif Mahmudah, Mesfin Kassaye Tessma, Yodi Mahendradhata

## Abstract

Multimorbidity has been increasing globally and is usually associated with higher health care utilization and costs. Indonesia has been implementing Universal Health Coverage (UHC) program since 2014. However, there is a limited study of the prevalence of multimorbidity and its impact on health care utilization and costs through the UHC scheme in Indonesia. This study aimed to determine the prevalence of multimorbidity and its associated factors, particularly the health care utilization and costs of patients with multimorbidity compared to patients with one chronic condition in the hospitals in Indonesia based on the UHC scheme. The study was a comparative cross-sectional design. The data was collected from the Social Security Agency for Health/Badan Penyelanggara Jaminan Sosial Kesehatan (BPJS Health) National Sample Data 2015-2016. All patients ≥60 years of age and have at least one chronic condition in the hospital were included. Descriptive statistics, bivariate analysis, and multivariable regression analysis were conducted to analyze the data. In a sample of 23,460 patients, the prevalence of multimorbidity was 44.4% among patients with chronic conditions in the hospital. We observed significant difference in gender, marital status, and membership segmentations between patients with multimorbidity and one chronic condition (p<0.05). Health care utilization and costs were significantly higher in multimorbid patients (p<0.001). This positive association between multimorbidity and health care utilization (OR: 1.70, 95% CI: 1.61-1.79) and health care costs (unstandardized coefficient 0.483, 95% CI: 0.443-0.524, p<0.001) remains significant after adjusting for age and gender. The analysis found that the prevalence of multimorbidity was high and positively associated with higher health care utilization and costs through the UHC scheme in Indonesia. Therefore, health policymakers and healthcare professionals need to consider the burden of multimorbidity more when structuring health care.

## Introduction

Multimorbidity is a coexistence of two or more chronic conditions in a single individual [1,2]. Compared to patients without a chronic condition and patients with only one chronic condition, multimorbidity is usually associated with higher health care utilization and cost, including more frequent hospital admission, longer hospital stays, and a need for a larger number of different medical specialists during a year [3–5]. Currently, the prevalence of multimorbidity is increasing both in high-income countries (HICs) and low and middle-income countries (LMICs), which increases with age, including in Indonesia [4,6].

Indonesia has been implementing Universal Health Coverage (UHC) program since 2014. It is implemented by the Social Security Agency for Health (Badan Penyelenggara Jaminan Sosial Kesehatan/BPJS Health) and covers 84.2% of the population in 2020[7,8]. In 2018, BPJS Health reported that chronic conditions were the highest claimed health expenditure with total of 42,171,533 cases and IDR 8,806,699,514,190 (equivalent to 638,239,743.66 USD) in outpatient care of hospitals [9].

To reduce the high treatment and financial burden of patients with chronic conditions, it is crucial to analyze the prevalence, associated factors, particularly the health care utilization and costs in patients, both with multiple chronic conditions (multimorbidity) and one chronic condition in Indonesia [3]. However, there is a limited study of the prevalence of multimorbidity and its impact on health care utilization and costs through the UHC scheme in Indonesia. This study aimed to determine the prevalence of multimorbidity and its associated factors, particularly the health care utilization and costs of patients with multimorbidity compared to patients with one chronic condition in the hospitals in Indonesia based on the UHC scheme.

## Methods

This study used a comparative cross-sectional design that determined the prevalence of multimorbidity and compared the health care utilization and costs between patients with multimorbidity and one chronic condition. We conducted and reported our study following the STROBE guideline [10].

### Sample and data

The data were collected from the BPJS Health National Sample Data from 2015 – 2016 in hospitals [11]. BPJS Health National Sample Data is an open-source database that provides information about the representative of the members nationally at the individual level. It comprises the characteristics of BPJS Health members, health care utilization, diagnosis, and related costs. The BPJS Health National Sample Data were collected by stratified random sampling method from the database of all members per December 31st, 2016. Each case has a different weight based on the stratification to reduce the bias between sample and population [12].

In this study, we included all patients aged 60 years, and older who were diagnosed with at least one chronic condition in hospital providers. This age group was chosen considering the life expectancy and the high prevalence of multimorbidity from a previous study in Indonesia [13]. The hospital setting was chosen because it was the only setting that provided data with more than one diagnosis to classify the cases into a patient with multimorbidity or a patient with one chronic condition. In this study, 19 chronic conditions were included based on the multimorbidity prevalence in Indonesia and ICD-10 codes used in the previous study [14,15] (Table 1).

**Table 1.**
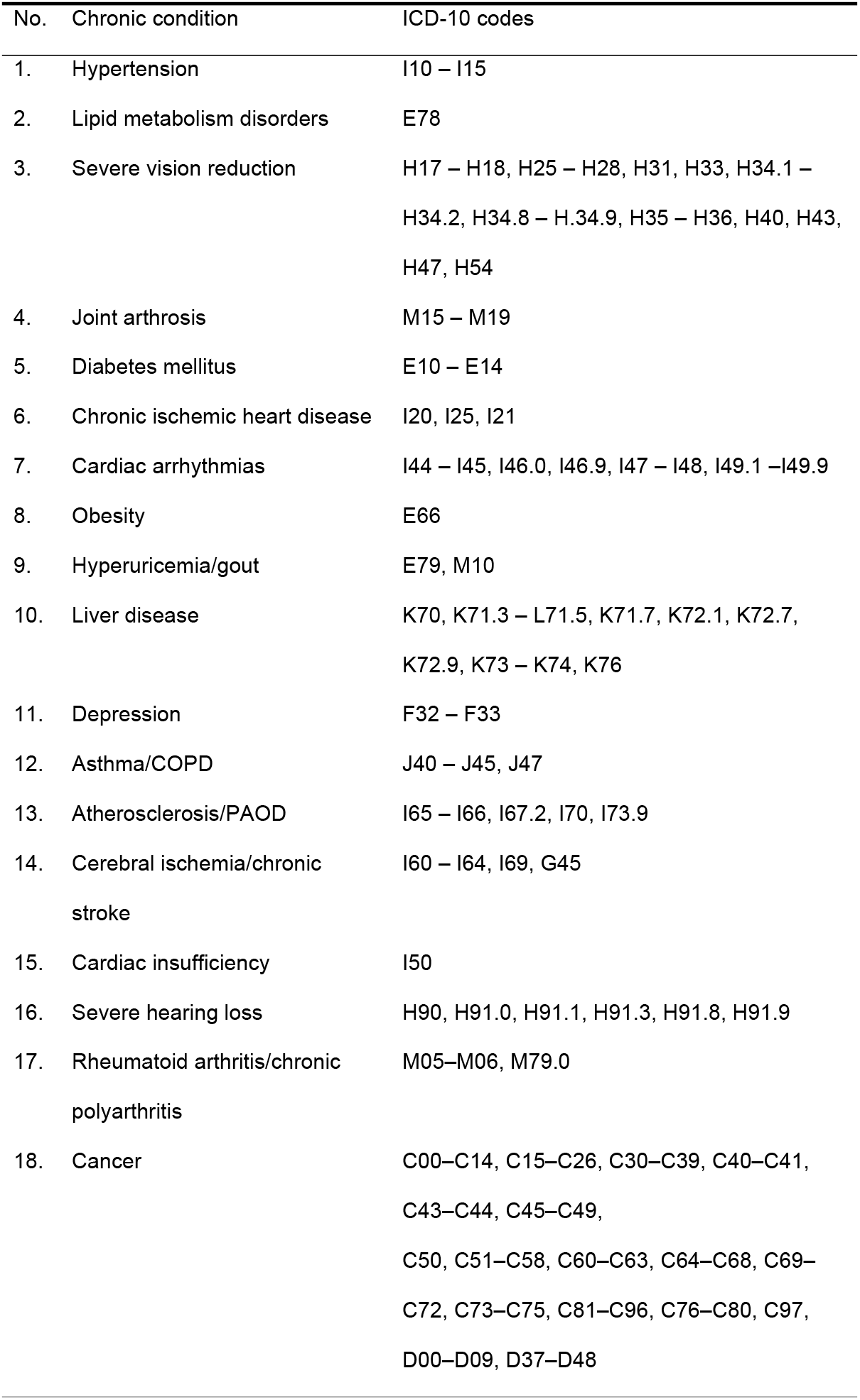
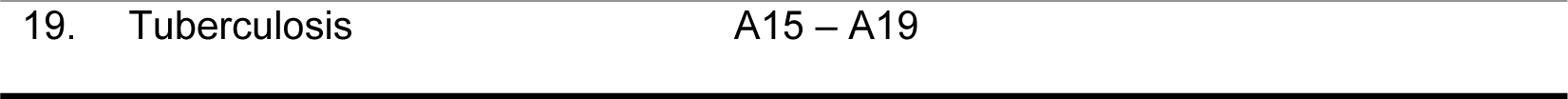
List of 19 chronic conditions based on ICD-10 codes.

Twenty-three thousand four hundred sixty subjects in the database that meet the inclusion criteria in the study were identified and included in this study (Fig 1). Subjects were then classified into with one chronic disease and with multimorbidity, two or more chronic diseases.

**Fig 1.**
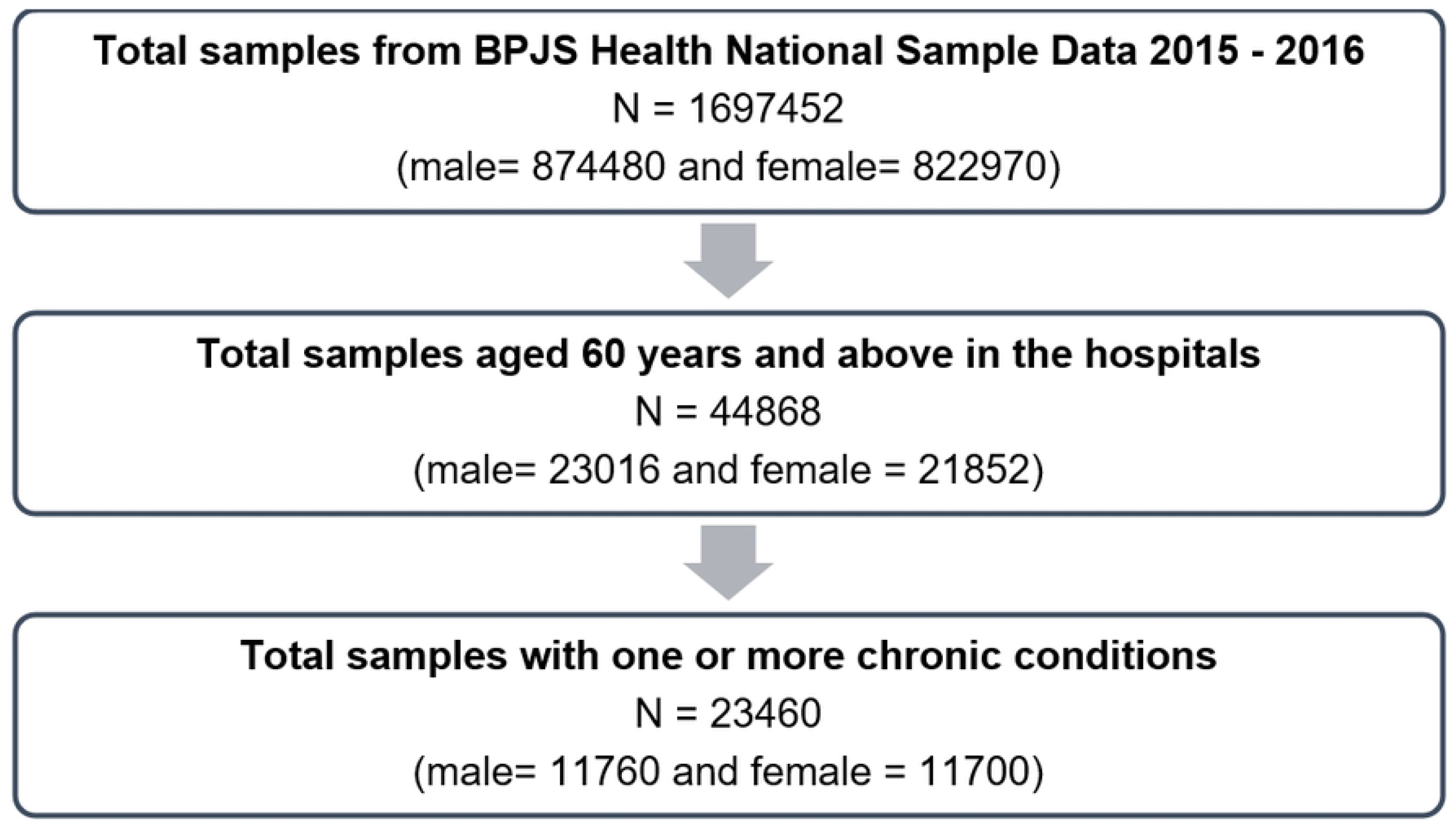
Selection of study sample from BPJS Health National Sample Data.

### Variables

#### Sociodemographic and Illness characteristics

The sociodemographic characteristics are determined by age group, gender, marital status, region, and membership segmentation. Membership segmentation of BPJS Health was classified into Premium Assistance Beneficiaries/*Penerima Bantuan Iuran* (PBI), who receives the assistance of contribution to BPJS health from government; and Non-Premium Assistance Beneficiaries/Non-Penerima Bantuan Iuran (Non-PBI), whose premium to BPJS Health is paid by the employers with contribution from employees, or themselves, or government (for the pensioners and veterans). Meanwhile, the illness characteristics are the severity level of diseases and the group of chronic conditions. The severity level of diseases ranged from level 0-3, with level 0 being the mildest, and level 3 being the most severe. Whereas the group of chronic conditions is based on the primary diagnosis of patients: cardiovascular, respiratory, special senses, cancer, endocrine, musculoskeletal, mental health [9,12,16].

#### Outcome variables

Health care utilization and costs were determined as the primary outcome variables. Health care utilization is a description of the use of health care services by persons to prevent and cure health problems [17]. This study includes the type of service (inpatient, outpatient), type of hospital (type A, type B, type C, type D, others), and length of stay.

The health care costs were according to the mix case-based group tariff rates, namely Indonesia Case-Based Groups (INA-CBGs), which follow the code of disease, severity level of disease, type of hospitalized room, and the number of secondary diagnosis[18,19]. The costing of INA-CBGs tariff rates was based on the overhead cost, intermediate cost, and final cost. The overhead cost is the cost for general hospital support, for example, the administration, maintenance, communication, cleaning service, information system, and nutrition. The intermediate cost is the medical hospital support, for instance, the pharmacy, radiology, laboratories, Intensive Care Unit (ICU), and hemodialysis. Meanwhile, the final cost is the medical care cost, including the outpatient and inpatient care [18,19]. The period for the cost for each individual is 2015 - 2016. The prices of INA-CBGs costs has been set by Ministry of Health, Republic of Indonesia in 2013 [19,20].

### Statistical analysis

Differences in patient characteristics between multimorbidity and one chronic condition were examined using a chi-square or a Fischer’s Exact test for categorical variables and a t-test for continuous variables. However, Mann-Whitney was performed for ordinal variables and in the case of a deviation from normality. For the ordinal variable level of exposure, we employed extended Mantel-Haenzel Chi-square for linear trend.

Multivariable regression was employed to control potential confounders and examine the relationship between the outcome and explanatory variables. Multiple linear regression was used to estimate the relationship between the continuous outcome variable (health care costs) and multimorbidity while controlling for other predictor or explanatory variables. Log transformation was applied to health care costs due to its high skewness. Multiple logistic regression was performed to examine the associations between the multimorbidity and demographic and other relevant explanatory variables. Explanatory variables were selected based on earlier research and univariable simple regression. The crude association of each explanatory variable was determined to examine its relationship with the outcome variable in univariate models.

Upon completing the univariate analyses, we selected variables for the multivariable analyses. Any variable whose univariate test had a P-value <0.15 was considered a candidate for the multivariable model along with variables of known practical importance. Once the variables were identified, they were entered into a multivariable model. Normal probability plots and residual plots were performed to assess violations of assumptions, and a Cook’s distance was examined to assess outlier problem. All analyses were conducted using IBM SPSS Statistic version 25.

## Results

### Sociodemographic and Illness characteristic of patients with multimorbidity versus one chronic condition

The total sample consisted of 23,460 patients with age equal to or more than 60 years and one or more medically diagnosed chronic conditions in the hospital providers. Of those individuals, 55.6% were patients with one chronic condition (n=13,034) and 44.4% were patients with multimorbidity (n=10,426). The mean (SD) age of the total sample was 70.6 (± 8.1), with 70.6 (± 8.1) for the patients with one chronic condition and 70.8 (± 8.0) for the multimorbidity patients.

Descriptive statistics of the sociodemographic and Illness characteristics of multimorbidity versus one chronic condition patients are shown in Table 2. We observed a statistically significant difference in some sociodemographic characteristics: gender, marital status, and membership segmentation.

**Table 2.**
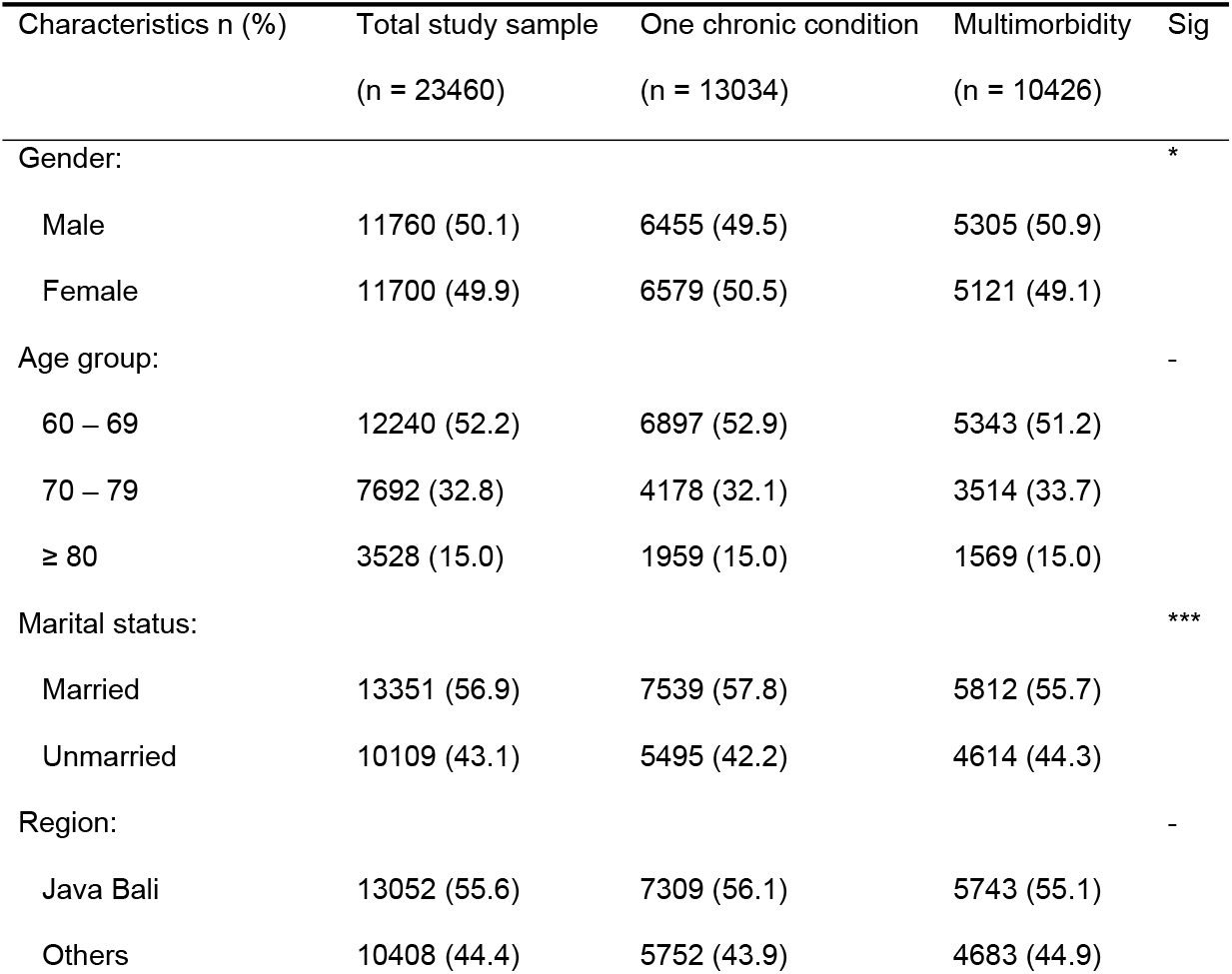
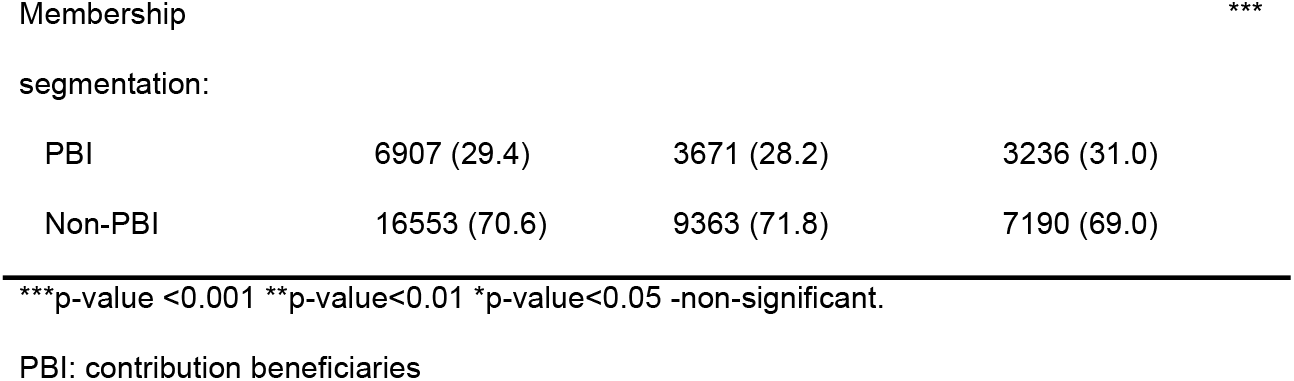
Sociodemographic characteristics of population aged ≥ 60 years with one chronic condition and multimorbidity in hospitals.

A significant difference was not observed in the age and region of patients with one chronic condition and multimorbidity. However, there was an increase in odds ratio among the age groups in multimorbidity (age group 60-69 as reference, OR=0.967, CI 95%:0.897-1.043 in the age group of 70-79, OR=1.05, CI 95%: 0.969-1.138 in age group ≥80).

Table 3 demonstrated illness characteristics. For the severity level of Illness, patients with one chronic condition had a higher proportion of level 0 compared to multimorbid patients. However, at levels 1-3, the proportion was higher in multimorbid patients compared to patients with one chronic condition. Besides, for the prevalence of chronic conditions, the three most frequent groups of chronic conditions were cardiovascular, respiratory, and endocrine. In contrast, the least frequent was mental health.

**Table 3.**
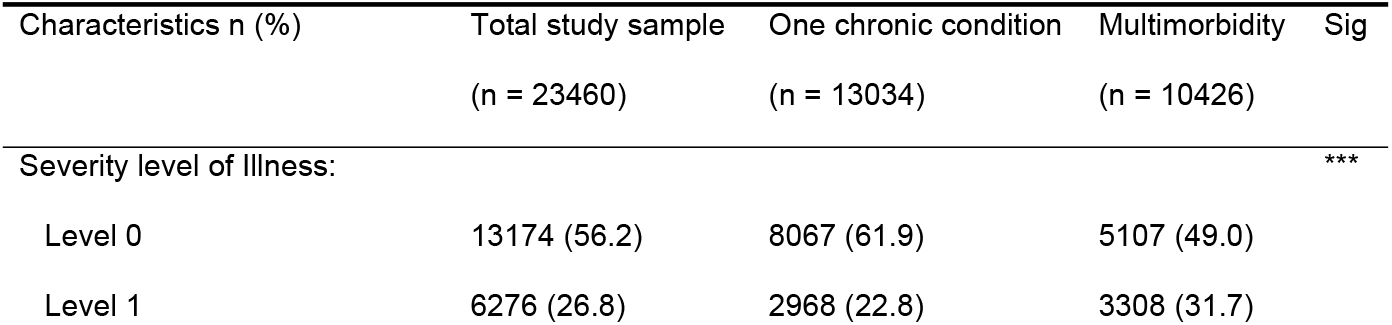
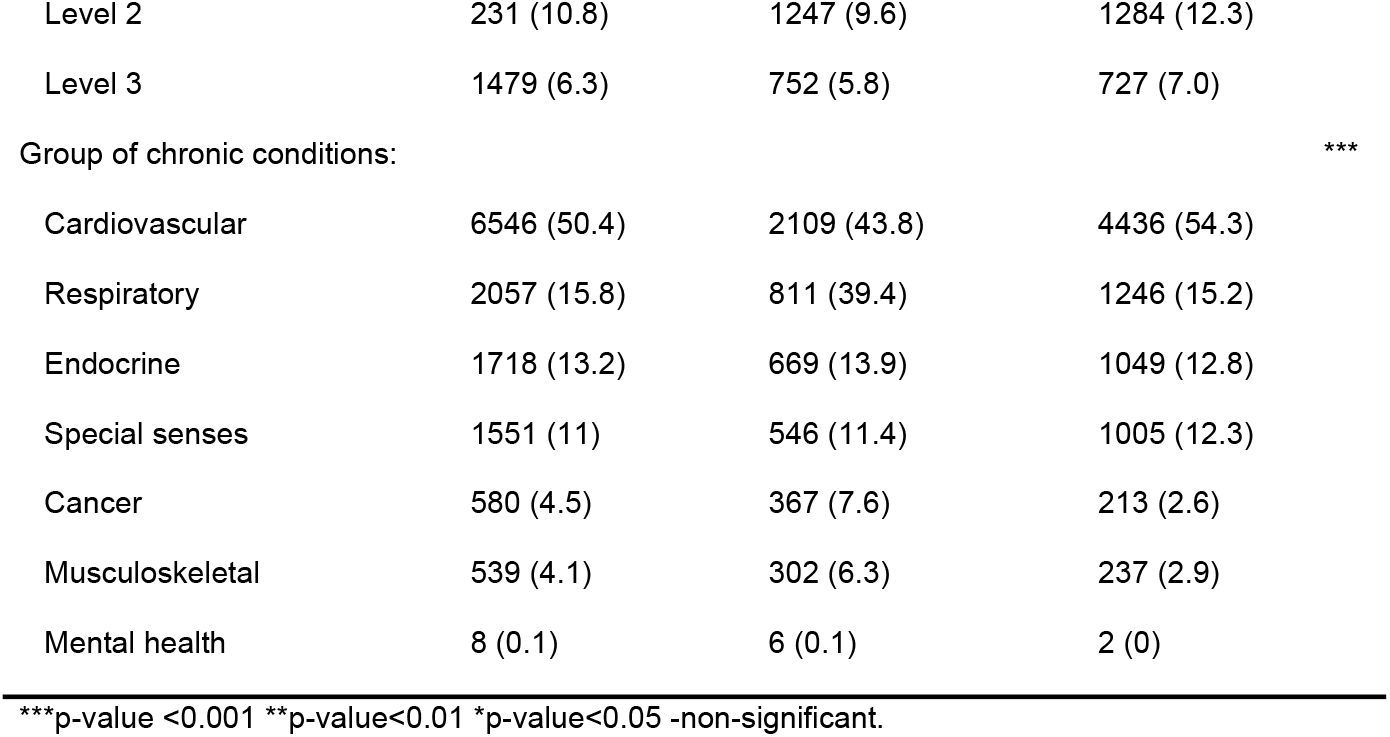
Illness characteristics of population aged ≥ 60 years with one chronic condition and multimorbidity in hospitals.

### Health care utilization and costs of patients with multimorbidity versus one chronic condition

As Table 4 shows, the proportion of patients admitted to the hospital (inpatient) was higher among patients with multimorbidity than patients with one chronic condition. The length of stay also was higher in patients with multimorbidity than patients with one chronic condition. However, the proportion of hospital type in patients with one chronic condition was similar to patients with multimorbidity in which the majority utilize the type C hospitals. The mean length of stay in days was 5.05 and 5.25 for patients with one chronic condition and patients with multimorbidity, respectively.

**Table 4.** Health care utilization and costs in population aged ≥ 60 years with one chronic condition and multimorbidity in hospitals, 2015-2016.

| Health care utilization | Total study sample | One chronic condition | Multimorbidity | Sig |
| --- | --- | --- | --- | --- |
| n (%) | (n = 23460) | (n = 13034) | (n = 10426) |  |
| Type of service: |  |  |  | *** |
| Inpatient | 10286 (43.8) | 4967 (38.1) | 5319 (51.0) |  |
| Outpatient | 13174 (56.2) | 8067 (61.9) | 5107 (49.0) |  |
| Type of hospital: |  |  |  | *** |
| Type A | 1209 (5.2) | 770 (5.9) | 439 (4.2) |  |
| Type B | 8331 (35.5) | 4681 (35.9) | 3650 (35.0) |  |
| Type C | 10333 (44.0) | 5741 (44.0) | 4592 (44.0) |  |
| Type D | 2597 (11.1) | 132 (10.1) | 1276 (12.1) |  |
| Others | 990 (4.2) | 521 (4.0) | 469 (4.5) |  |
| Length of stay, days | 4 (4) | 4 (4) | 4 (5) | - |
| median (IQR) |  |  |  |  |
| Health care costs, IDR | 307 450.00 | 190 900.00 | 2 160 050.00 | *** |
| median (IQR) | (3 552 200) | (3 034 175) | (4 117 800) |  |

\*\*\*p-value <0.001 \*\*p-value<0.01 \*p-value<0.05 -non-significant. IDR: Indonesian Rupiah,
IQR: interquartile

For the health care costs, multimorbid patients also had higher costs than patients with one chronic condition (Table 4). The average health care costs in patients with one chronic condition was IDR 2,122,912.19 per year (equal to USD 140.47). While the average health care costs in patients with multimorbidity was IDR 2,884,995.96 per year (equal to USD 190.90).

### Underlying factors associated with health care utilization and costs of patients with multimorbidity

In the multiple logistic regression model on health care utilization, multimorbidity was positively associated with inpatient care. The odds ratio (OR) in multimorbidity for inpatient care remains statistically significant, and the association positive when corrected for gender and age. Thus, adjusting for gender and age, the inpatient costs is higher for multimorbid patients. Table 5 shows no remarkable difference in the unadjusted and adjusted OR.

**Table 5.** Results of multiple logistic regression model on the dependent variable of inpatient health care utilization (yes/no) in population aged ≥ 60 years.

| Model | Unadjusted OR (95% CI) | Adjusted OR (95% CI) |
| --- | --- | --- |
| Multimorbidity | 1.69 (1.61-1.78) | 1.70 (1.61-1.79) |
| Age group | 1.47 (1.42-1.53) | 1.48 (1.43-1.53) |
| Gender | 0.92 (0.88-0.97) | 0.92 (0.87-0.97) |

OR: Odds ratio, CI: Confidence intervals

In addition, a multiple linear regression was performed to determine if the observed variations in health care costs were either attenuated or eliminated after adjusting for demographic confounding variables. The dependent variable was the log health care costs. After adjustment, statistically significant differences were observed in the variables multimorbidity, gender, and age group. Multimorbidity remained statistically significant after adjusting for gender and age group with higher Beta values. Thus, multimorbidity was observed to be a significant positive predictor of log health care costs, regression coefficient (B) = .48 (95% CI = .44, .52, P < 0.001). Female gender was negatively related to log health care costs, B = −0.07 (95% CI = −.11, -.03, P< 0.001). Age group was positively related to the dependent variable, B = .30. (95%CI= .27, .33, P<0.001).

The coefficient of multimorbidity indicated that for patient with multimorbidity, it was expected that the log costs increase by an average of 0.48. The coefficient of age group indicated that for increasing age group, the log costs are expected to increase by an average of 0.30 The coefficient of gender indicated that in female gender compared to male, the log costs expected to reduce by an average of 0.07. Hence, multimorbidity, gender, and age group were significant predictors of log health care costs (Table 6).

**Table 6.** Results of multiple linear regression model for the dependent variable log health care costs, in population aged ≥ 60 years.

| Predictor | Unstandardized |  | Standardized |  | t | Sig. | 95% Confidence |  |
| --- | --- | --- | --- | --- | --- | --- | --- | --- |
|  | Coefficients |  | Coefficients |  |  |  | Interval for B |  |
|  | B | Std. Error | Beta |  |  |  | Lower | Upper |
|  |  |  |  |  |  |  | Bound | Bound |
| Multimorbidity | .483 | .021 | .148 |  | 23.094 | *** | .443 | .524 |
| Gender | -.070 | .021 | .134 |  | 20.970 | *** | -.112 | -.030 |
| Age group | .298 | .014 | -.022 |  | -3.394 | *** | .270 | .326 |

\*\*\*p-value <0.001

## Discussion

### Principal findings

#### Prevalence of multimorbidity in Indonesia based on Indonesia UHC program

In the hospitals, half of the patients aged 60 years and above had at least one chronic condition, in which 44% had multimorbidity. This prevalence of multimorbidity was lower than the previous study in Indonesia by Hussain et al. [14], which stated that half of the individuals over 60 years in Indonesia had multimorbidity. This difference is possible due to the limited population in the hospitals in the current study rather than in all health care settings.

The sociodemographic characteristics which significantly different were gender, marital status, and membership segmentation. In contrast to majority studies, the males had a higher prevalence of multimorbidity than females in this study [5,14]. However, this reverse trend was also observed in other study [21]. The finding for marital status was in line with previous studies [22,23]. This proportion might be due to the high number of people that married among the population aged 60 years and above [24]. Additionally, the membership segmentation that reflected the socioeconomic situation of the patients was also in line with other studies [14,25]. A previous study in Indonesia [14] also showed that higher socioeconomic status is more vulnerable to multimorbidity as the epidemiological transition in LMICs. The wealthier stratum has a higher risk for Non-Communicable Diseases in LMICs with higher levels of health risk such as high cholesterol, waist-hip ratio, and reduced physical activity [22].

On the other hand, the mean of age, proportion of age group and region were not significantly different in multimorbid patients compared to the patients with one chronic condition. It might be due to the current epidemiological transition in Indonesia, which leads to chronic conditions in all ages and across the geographic area [26]. However, the prevalence of multimorbidity increased with age, which aligned with numerous studies [5,13,27].

The illness characteristics of severity level and group of chronic conditions were significantly different. In alignment with previous studies [22,28], the severity level of Illness showed that the multimorbidity had a higher proportion in more severe conditions. The negative effect on multimorbidity in elderly age groups has been reported from high- and low-income settings [4,27]. The prevalence of a group of chronic conditions confirms some of the findings from a previous study in Indonesia [14], including that the most frequent chronic conditions group was cardiovascular.

#### Health care utilization and costs of patients with multimorbidity and with one chronic condition

The results showed that health care utilization and costs were higher in patients with multimorbidity than patients with one chronic condition. The inpatient hospital admission was higher in multimorbid patients. Moreover, the length of stay was also longer compared to patients with one chronic condition. These findings confirm that multimorbidity is related to higher health care utilization [4,5,29,30]. A cross-sectional study in LMICs showed that visits to doctors in secondary care and hospitalization increased with multimorbidity [31].

The utilization of different types of hospital was also significantly different between patients with multimorbidity and one chronic condition. Nevertheless, the proportion of hospital type in patients with one chronic condition was similar to patients with multimorbidity in which the majority utilize the type C hospitals. This proportion is possibly due to the tiered referral system of BPJS Health from primary care to type D or C hospitals, then to type B, and type A as top referral [16].

Health care costs were higher in patients with multimorbidity than with one chronic condition, which is similar to other studies [27,30,32]. For instance, a recent systematic review study, which included studies from the United States, Europe, Canada, stated that there was a positive correlation between multimorbidity and health care costs [27]. Glynn et al. also reported that health care costs were significantly increased among multimorbid patients [27].

#### Underlying factors associated with health care utilization and costs of patients with multimorbidity

This study found a positive association between the utilization of inpatient health services and multimorbidity. This association remains positive and increased to 70% after adjusting for gender and age. Age group and gender also significantly associated with health care utilization. In contrast, the female gender had negative association to the type of services.

Those findings were in line with several previous studies [27,32,33], both in HICs and LMICs. Studies in high-income countries found that hospital admission was significantly increased with an increase in the number of chronic conditions after adjusting for age, gender, and free medical care eligibility [27,32,34]. A study in low-middle income countries found that the increased number of chronic conditions was significantly associated with higher health care utilization, including hospitalization and duration of stay in hospital [35].

Multimorbidity also had a positive association with health care costs. The result showed that multimorbidity was a positive predictor of health costs in alignment with other research findings [35]. The increased costs may be related to the associated costs of caring for individuals with multimorbidity problems due to increased medical expenditures. Thus, multimorbidity also incurs significant social and economic burdens due to complex health and welfare demands and higher health costs.

Increasing age group also was shown to be a positive predictor. Consistent with the previous study which have found that this increase in health care costs is reasonable considering the ageing population with multimorbidity [36]. However, female gender was a negative predictor. A recent study also reported that the combination of diseases and intensity of multimorbidity differed in older men and women [36]. Further, several studies also found that the number of chronic conditions was associated with higher total health care costs after controlling for age and gender, severity of Illness, and hospital characteristics [37–39].

### Strength and limitation

To the best to our knowledge, this study is the first to determine the multimorbidity and its associated factors in the Indonesian population with an age equal to or more than 60 years using data from BPJS Health National sample data. A major strength of the current study was the large stratified randomly selected representative sample of the Indonesian population through BPJS Health. Thus, minimizing selection bias and enables for larger generalization of results. Nevertheless, there are some study limitations.

The study was cross-sectional, which did not allow to determine any causal interferences. Our findings may have underestimated the prevalence of multimorbidity since the scope was limited to hospital settings and based on the claims data. Nonetheless, the diagnosis of chronic conditions was ensured following the ICD-10 diagnosis rather than self-report. The current study may also have underestimated the health care utilization and costs of multimorbidity. The data only provided the total health care costs based on INA-CBGs rather than in specific detail of each unit of costs. This limited data made the study unable to examine the health care costs of multimorbidity. Lastly, the administrative claims also did not record more details in economic and social factors such as income and education levels that might independently influence the health care utilization and costs.

### Implications to Health Care System

The prevalence of multimorbidity among patients with chronic conditions and the positive association between multimorbidity with higher health care utilization and costs in BPJS Health should urgently be addressed by health care professionals and policy makers in Indonesia.

Multimorbidity is a complex disease, and its management needs to be improved [5,40]. However, the current administration has been focused on single conditions approach rather than multiple conditions [5,40,41]. Therefore, cooperation, coordination, and communication between different levels of health care and specialties need to be increased. It is also crucial to develop an integrated care pathway and guideline to manage the multimorbidity. Further, cost-effective management of multimorbidity and their risk factors should also be improved [5].

Due to the high burden of multimorbidity in BPJS Health, the health policymakers and health care professionals should promote a healthy lifestyle to prevent the risk factors. BPJS Health should also expand the coverage to preventive measures than focus only on curative to avoid the high burden of health financing, particularly multimorbidity in the future [16,26].

The findings showed the association and impact of multimorbidity in health care utilization and costs, particularly in Indonesia through UHC scheme. However, further research is needed to understand the pattern of chronic conditions in multimorbidity. Cluster analysis by grouping similar diseases may improve the knowledge of how multimorbidity as complex disease impact health care utilization and costs. A study with more comprehensive data from different health care levels, including sociodemographic of respondents, could provide further context for multimorbidity research.

## Conclusions

This study is a step forward to examine the impact of multimorbidity on health care utilization and costs in Indonesia through UHC scheme. The study found a 44.4% prevalence of multimorbidity among patients with chronic conditions in hospitals. Multimorbidity also had a positive association with higher health care utilization and costs compared to patients with one chronic condition. Therefore, health policymakers and health care professionals need to take the burden of multimorbidity more into consideration when structuring health care.

Further research could be directed toward the pattern of chronic conditions in multimorbidity with more comprehensive data from different levels of health care.

## Data Availability

Data are available from BPJS Health (https://data.bpjs-kesehatan.go.id/). To gain access to BPJS Health sample data, the researchers need to submit an application to the BPJS Health Information and Documentation Management Officer. This is to ensure the data are properly monitored and in accordance to data governance-based policy.

## Acknowledgement

The authors express sincere gratitude to BPJS Health for providing access to the data.

